# Post-Anticoagulant D-dimer as a Highly Prognostic Biomarker of COVID-19 Mortality

**DOI:** 10.1101/2020.09.02.20180984

**Authors:** Xiaoyu Song, Jiayi Ji, Boris Reva, Himanshu Joshi, Anna Pamela Calinawan, Madhu Mazumdar, Emanuela Taioli, Pei Wang, Rajwanth Veluswamy

## Abstract

**Importance:** Clinical biomarkers that accurately predict mortality are needed for the effective management of patients with severe COVID-19 illness.

**Objective:** To determine whether D-dimer levels after anticoagulation treatment is predictive of in-hospital mortality.

**Design:** Retrospective study using electronic health record data.

**Setting:** A large New York City hospital network serving a diverse, urban patient population.

**Participants:** Adult patients hospitalized for severe COVID-19 infection who received therapeutic anticoagulation for thromboprophylaxis between February 25, 2020 and May 31, 2020.

**Exposures:** Mean and trend of D-dimer levels in the 3 days following the first therapeutic dose of anticoagulation.

**Main Outcomes:** In-hospital mortality versus discharge.

**Results:** 1835 adult patients (median age, 67 years [interquartile range, 57-78]; 58% male) with PCR-confirmed COVID-19 who received therapeutic anticoagulation during hospitalization were included. 74% (1365) of patients were discharged and 26% (430) died in hospital. The study cohort was divided into four groups based on the mean D-dimer levels and its trend following anticoagulation initiation, with significantly different in-hospital mortality rates (p<0.001): 49% for the high mean-increase trend (HI) group; 27% for the high-decrease (HD) group; 21% for the low-increase (LI) group; and 9% for the low-decrease (LD) group. Using penalized logistic regression models to simultaneously analyze 67 variables (baseline demographics, comorbidities, vital signs, laboratory values, D-dimer levels), post-anticoagulant D-dimer groups had the highest adjusted odds ratios (ORadj) for predicting in-hospital mortality. The ORadj of in-hospital death among patients from the HI group was 6.58 folds (95% CI 3.81-11.16) higher compared to the LD group. The LI (ORadj: 4.06, 95% CI 2.23-7.38) and HD (ORadj: 2.37; 95% CI 1.37-4.09) groups were also associated with higher mortality compared to the LD group.

**Conclusions and Relevance:** D-dimer levels and its trend following the initiation of anticoagulation have high and independent predictive value for in-hospital mortality. This novel prognostic biomarker should be incorporated into management protocols to guide resource allocation and prospective studies for emerging treatments in hospitalized COVID-19 patients.

**Key Points:** *Question:* Are D-dimer levels following therapeutic anticoagulation predictive of mortality in hospitalized COVID-19 patients?

*Finding:* In a retrospective study of 1835 adult COVID-19 patients who received therapeutic anticoagulation for thromboprophylaxis during hospitalization, 1365 (74%) patients were discharged and 470 (26%) died. Post-anticoagulant D-dimer levels and trends were significantly and independently predictive of mortality.

*Meaning:* Active monitoring of post-anticoagulant D-dimer levels in hospitalized COVID-19 patients is a novel strategy for stratifying individual risk of in-hospital mortality that can help guide resource allocation and prospective studies for emerging treatments for severe COVID-19 illness.

## Introduction

The COVID-19 pandemic, caused by the viral pathogen severe acute respiratory syndrome coronavirus 2 (SARS-CoV-2), has resulted in over than 22.2 million confirmed cases and 783,000 deaths worldwide through August, 2020 ^1^. Among patients with more severe illness requiring hospitalization, there is an urgent need for accurate clinical biomarkers to predict mortality risk in order to guide clinical decisions, allocate critical resources and inform study designs of emerging treatments. Currently, there is insufficient evidence to precisely identify patients at the highest risk of poor outcomes, and clinicians often consider a multitude of individual clinical factors (i.e., exam findings, laboratory tests) without predictive cut-off values for making treatment decisions. Therefore, quantifying the impact of individual clinical factors and development of novel prognostic biomarkers for clinical outcomes is critical for the effective management of COVID-19 patients.

D-dimer, a small protein fragment present in blood resulting from plasmin cleavage of cross-linked fibrin clots, is routinely used in clinical practice as a sensitive biomarker in the evaluation of venous thromboembolism (VTE) ^2^. Recently, several studies have shown that elevated D-dimer levels at the time of hospital admission in COVID-19 patients are associated with higher mortality, sparking significant interest in understanding the role of D-dimer in these patients ^3–6^. D-dimer levels reflect the underlying hypercoagulable state in COVID-19 patients, and the use of anticoagulant therapy in hospitalized COVID-19 patients with elevated D-dimer levels resulted in a significant mortality benefit ^3,7–10^. As a consequence, many guidelines and institutional protocols have recommended therapeutic (using intermediate or full doses of anticoagulants) anticoagulation strategies for thromboprophylaxis in patients with severe COVID-19 infection, particularly for those having significantly elevated baseline D-dimer levels ^4–6^. However, while D-dimer measurements generally are followed throughout the hospitalization, there remains no consensus or guidance as to how D-dimer levels should be monitored or interpreted with respect to anticoagulant therapy and outcomes in COVID-19 patients.

In this study, we hypothesized that the D-dimer levels and their trends following therapeutic anticoagulation in patients with severe COVID-19 infections may be predictive of mortality in addition to other known risk factors. To determine the role of D-dimer in this setting, we leveraged a large institutional database of COVID-19 hospitalized patients from the Mount Sinai Health System (MSHS) in New York City, one of the initial epicenters of the COVID-19 pandemic in the United States (US).

## Methods

### Study Cohort

MSHS includes the Mount Sinai Hospital and 7 other tertiary and community hospitals throughout New York City, serving a diverse patient population with a high representation of low-income minorities. This study utilized a comprehensive COVID-19 database created by the Mount Sinai Data Warehouse, which includes de-identified clinical data extracted from the electronic medical records of all patients tested for and/or diagnosed with COVID-19 until May 31, 2020. We included all adult (≥18 years of age) patients who were hospitalized for a new COVID-19 infection (based on RT-PCR COVID-19 assay using nasopharyngeal swabs), and were treated with therapeutic anticoagulation for thromboprophylaxis. Of those, patients having follow up data for at least 3 days after the first anticoagulant dose and information on their hospitalization outcome (discharged vs. deceased) were included. We then excluded patients who: 1) had absolute contraindications for therapeutic anticoagulation due to either low platelet counts (<50,000/uL) or elevated international normalization ratio (INR>1.5); 2) were given therapeutic anticoagulation or tissue plasminogen activator (TPA) for a newly diagnosed VTE as large vessel thrombosis could affect post-anticoagulant D-dimer levels; and 3) were discharged and readmitted into the hospital as baseline information for each admission could not be uniquely identified.

### Study Variables

#### Patient Characteristics

For each patient, we obtained baseline sociodemographic data (i.e., age, sex, self-reported race and ethnicity), smoking status, body mass index (BMI), and 18 common comorbidities (**Supplemental Table 1**). Baseline vital signs (temperature, systolic blood pressure, diastolic blood pressure, oxygen saturation, heart rate, respiratory rate) and laboratory tests obtained within 24 hours of admission and prior to receiving anticoagulation were collected. In cases where multiple vital signs were recorded during the first 24 hours of admission, we used the most clinically abnormal measurement concerning for systemic inflammatory response syndrome (SIRS) ^11^. A preprocessing procedure was performed to exclude laboratory tests that were missing in > 50% of patients. The remaining 35 laboratory tests that were used for analysis included complete blood count (CBC) with differential, complete metabolic panel (CMP), inflammatory markers (i.e., ferritin, C-reactive protein [CRP], Lactate dehydrogenase [LDH]) liver function tests and baseline D-dimer. All together, 65 baseline variables were considered for each patient (**Supplemental Table 1**).

#### Therapeutic dose of anti-coagulation treatment

The MSHS, alongside many other high acuity hospitals, has developed a standardized protocol for anticoagulant therapy in patients requiring hospital admission for COVID-19 and without increased risk of bleeding. All patients are assessed for VTEs and if confirmed or had a high level of clinical suspicion, treatment-dose anticoagulation is recommended. All other patients are recommended to receive thromboprophylaxis with heparin, enoxaparin and/or apixaban using either prophylactic doses for patients without severe respiratory compromise or therapeutic doses (intermediate or full) for patients with severe respiratory compromise. Our study cohort included patients on therapeutic doses defined as: 1) heparin: >5,000 units subcutaneous (SQ) every 8 hours in patients with BMI<40 kg/m2 or >7,500 units SQ every 8 hours in patients with BMI ≥40 kg/m2; 2) enoxaparin: 1mg/kg SQ every 24 hours (intermediate dose) or 1mg/kg SQ twice daily (full dose); and 3) apixaban: >2.5mg by mouth every 24 hours.

#### Post-anticoagulant D-dimer values and groups

We recorded post-anticoagulant D-dimer levels as all measurements collected within the first 3 days after therapeutic anticoagulation was started. As the number of D-dimer measurements during this period varied dramatically (from 0 to 18 measurements) for each patient, we calculated both the mean and trend to summarize the data. The trend was defined, if at least 2 measurements were available, as the slope of a linear regression model characterizing the dependence of the post-anticoagulant D-dimer values on the test collection time from anticoagulation.

Using 2.5 ug/ml as a cutoff for the post-anticoagulant D-dimer mean value, and 0 as a cutoff for the post-anticoagulant D-dimer trend, we divided patients into four groups with similar sample sizes: HI--- high mean value (>=2.5 ug/ml) and increase trend (trend>=0); HD--- high mean value and decrease trend; LI --- low mean value and increase trend; and LD --- low mean value and decrease trend.

### Study End Point

The study endpoint is a binary indicator of in-hospital mortality, defined as patients who died during their admission vs. patients who were discharged alive from the hospital, usually to home, nursing facility, acute/sub-acute rehab or long-term care facility.

#### Statistical Analysis

To test the associations between baseline and post-anticoagulant D-dimer variables with in-hospital mortality, *χ^2^* tests and two-sample Wilcoxon tests were used for categorical and continuous variables, respectively. Bonferroni correction for multiple testing provided p<0.001 (=0.1/65) as the cutoff to determine significant associations with in-hospital mortality. Missing values in categorical variables were treated as a separate category, while multiple imputations were performed for missing values in numeric variables using the R package MICE^12^.

Logistic regression models were employed to predict in-hospital mortality based on baseline and post-anticoagulant D-dimer levels. The predictive values were evaluated through 10-fold cross-validation. The Receiver Operating Characteristic (ROC) curve and the corresponding AUCs (Area under the ROC Curves) were used to assess and compare the performance of the prediction model based on baseline and post-anticoagulant D-dimer values.

We assessed whether the in-hospital mortality and the baseline characteristics of patients differed across the four D-dimer groups described above. *χ^2^* tests were used for categorical variables and Kruskal-Wallis tests for continuous variables. Variables that passed 5% significance level after Bonferroni correction were further examined for statistically significant differences in groups with high D-dimer levels (HI and HD combined) vs. groups with low D-dimer levels (LI and LD combined); as well as in groups with increasing D-dimer vs. those with decreasing D-dimer trends (HI vs. HD; LI vs. LD).

We assessed the predictivity of D-dimers for in-hospital mortality conditional on baseline characteristics of patients. To better estimate effect sizes of predictors, we randomly split the samples into a discovery and validation subset with equal sizes and performed variable selection on the discovery subset while inference of effect sizes on the validation subset to avoid post-selection inference, which results in biased estimates and confidence intervals [CIs]. In the discovery subset, we utilized regularized logistic regression models with Lasso penalty^13^ to select the most important predictors for in-hospital mortality from a large feature set including the indicators of the post-anticoagulant D-Dimer groups and 65 baseline variables (**Supplementary Table 1**). For the variables selected by the penalized regressions, we performed an ordinary logistic regression using the validation subset to estimate odds ratios (ORs) and the corresponding 95% CIs. For the variables that confirmed to be statistically significant in the validation subset, we calculated AUC differences between leave-one-predictor-out models and the full model to assess the relative importance of each predictor. Moreover, we performed parallel analyses using only baseline variables, and compared the predictive performance of these models with the above ones using post-anticoagulant D-Dimer information. We compared the 10-fold cross-validation prediction AUCs between the baseline model and the full model with post-anticoagulant D-dimer groups among 100 randomly selected bootstrap samples of validation subset. All statistical analyses were repeated in complete case analysis among samples without missing post-anticoagulant D-dimer data, with similar results (**Supplementary Table 3**).

## Results

### Baseline Characteristics of Study Cohort

After applying the selection criteria to the COVID-19 database (n=65,501 patients), the final study cohort consisted of 1835 laboratory-confirmed COVID-19 positive adult patients who were hospitalized in the MSHS between February 25 and May 31, 2020 (**Figure 1**). Among them, 470 (26%) study patients died during hospitalization and 1365 (74%) were discharged alive. Patients who died during hospitalization were generally older, had more comorbidities, presented with signs of more severe respiratory distress (higher respiratory rates and lower minimum oxygen saturation), had lower kidney function, higher levels of inflammatory markers (ferritin, CRP, LDH) and higher baseline D-dimers (p<0.001 for all comparisons after Bonferroni correction). Although not statistically significant, patients who died during hospitalization also experienced a longer time between admission and the start of therapeutic anticoagulation (**Tables 1 and S1**).

**Table 1.** Characteristics of Hospitalized COVID-19 Patients According to Survival Status.

| Characteristics | Discharged from Hospital<br>(N=1365) | Deceased in Hospital<br>(N=470) | P-value |
| --- | --- | --- | --- |
| Age (years), median (IQR<sup>1</sup>) | 65 [54, 75] | 73 [65, 83] | <0.001* |
| Male sex, N (%) | 805 (59.0) | 262 (55.7) | 0.242 |
| Race/Ethnicity, N (%) |  |  |  |
| White | 275 (20.1) | 111 (23.6) | 0.374 |
| Black | 277 (20.3) | 83 (17.7) |  |
| Hispanic | 404 (29.6) | 138 (29.4) |  |
| Other | 236 (17.3) | 73 (15.5) |  |
| Missing | 173 (12.7) | 65 (13.8) |  |
| Smoking Status, N (%) |  |  |  |
| Never smoker | 688 (50.4) | 229 (48.7) | 0.714 |
| Ever smoker | 384 (28.1) | 132 (28.1) |  |
| Missing | 293 (21.5) | 109 (23.2) |  |
| Baseline comorbidities, N (%) (Details in Table S1) |  |  |  |
| 0 | 456 (33.4) | 89 (18.9) | <0.001* |
| 1-2 | 534 (39.1) | 196 (41.7) |  |
| >2 | 375 (27.5) | 185 (39.4) |  |
| BMI<sup>2</sup> (kg/m<sup>2</sup>), median (IQR) | 27.55 [23.88, 32.12] | 27.46 [24.32, 32.48] | 0.331 |
| Admission Vital Signs, median (IQR) |  |  |  |
| Systolic Blood Pressure maximum (mmHg) | 143 [129, 159] | 144 [129, 161] | 0.531 |
| Diastolic Blood Pressure maximum (mmHg) | 83 [76, 92] | 81 [73, 91] | 0.017 |
| Heart Rate maximum (beats/minute) | 101.00 [90.75, 114.00] | 103.00 [91.00, 120.75] | 0.009 |
| Respiratory Rate maximum (breaths/minute) | 22 [20, 28] | 24 [20, 30] | <0.001* |
| Temperature maximum (Fahrenheit) | 99.8 [98.6, 101.3] | 99.6 [98.6, 101.1] | 0.064 |
| Oxygen Saturation minimum (%) | 93 [90, 95] | 90 [84, 94] | <0.001* |
| Baseline Laboratory Tests, median (IQR) |  |  |  |
| White Blood Cell (x10E3/uL) | 7.60 [5.50, 10.40] | 8.14 [5.90, 12.28] | 0.011 |
| Lymphocyte % | 13.05 [8.30, 18.58] | 11.00 [7.25, 16.95] | 0.003 |
| Neutrophil % | 78.60 [70.45, 84.77] | 81.90 [73.40, 87.10] | <0.001* |
| Hemoglobin (g/dL) | 12.70 [11.10, 13.90] | 12.90 [11.10, 14.10] | 0.448 |
| Platelet (x10E3/uL) | 221.00 [167.00, 293.00] | 187.00 [147.00, 244.00] | <0.001* |
| Serum Creatinine (mg/dL) | 0.95 [0.73, 1.41] | 1.20 [0.82, 2.00] | <0.001* |
| Estimated Glomerular Filtration Rate<br>(ml/min/1.73m<sup>2</sup>) | 54.00 [30.15, 77.00] | 41.28 [26.00, 58.33] | <0.001* |
| Alanine Aminotransferase (u/L) | 30.00 [19.00, 50.00] | 30.00 [20.25, 50.75] | 0.885 |
| Aspartate Aminotransferase (u/L) | 40.00 [27.00, 63.50] | 48.00 [33.00, 71.75] | <0.001* |
| Total Bilirubin (mg/dL) | 0.60 [0.40, 0.80] | 0.50 [0.40, 0.80] | 0.356 |
| C-Reactive Protein (mg/L) | 113.50 [53.70, 194.30] | 159.10 [90.82, 235.70] | <0.001* |
| Ferritin (ng/mL) | 697.00 [330.00, 1512.00] | 1038.00 [480.00, 2092.00] | <0.001* |
| Lactate Dehydrogenase (u/L) | 412.00 [312.00, 528.50] | 516.50 [381.25, 660.75] | <0.001* |
| Baseline D-dimer (ug/mL) | 1.37 [0.79, 2.46] | 1.76 [1.08, 2.92] | <0.001* |
| Days from admission to start of anticoagulation,<br>median (IQR) | 0.55 [0.20, 1.68] | 0.67 [0.24, 2.13] | 0.017 |
| <sup>1</sup> Interquartile Range; <sup>2</sup>Body Mass Index; * Significant after multiple testing |  |  |  |

**Figure 1.**
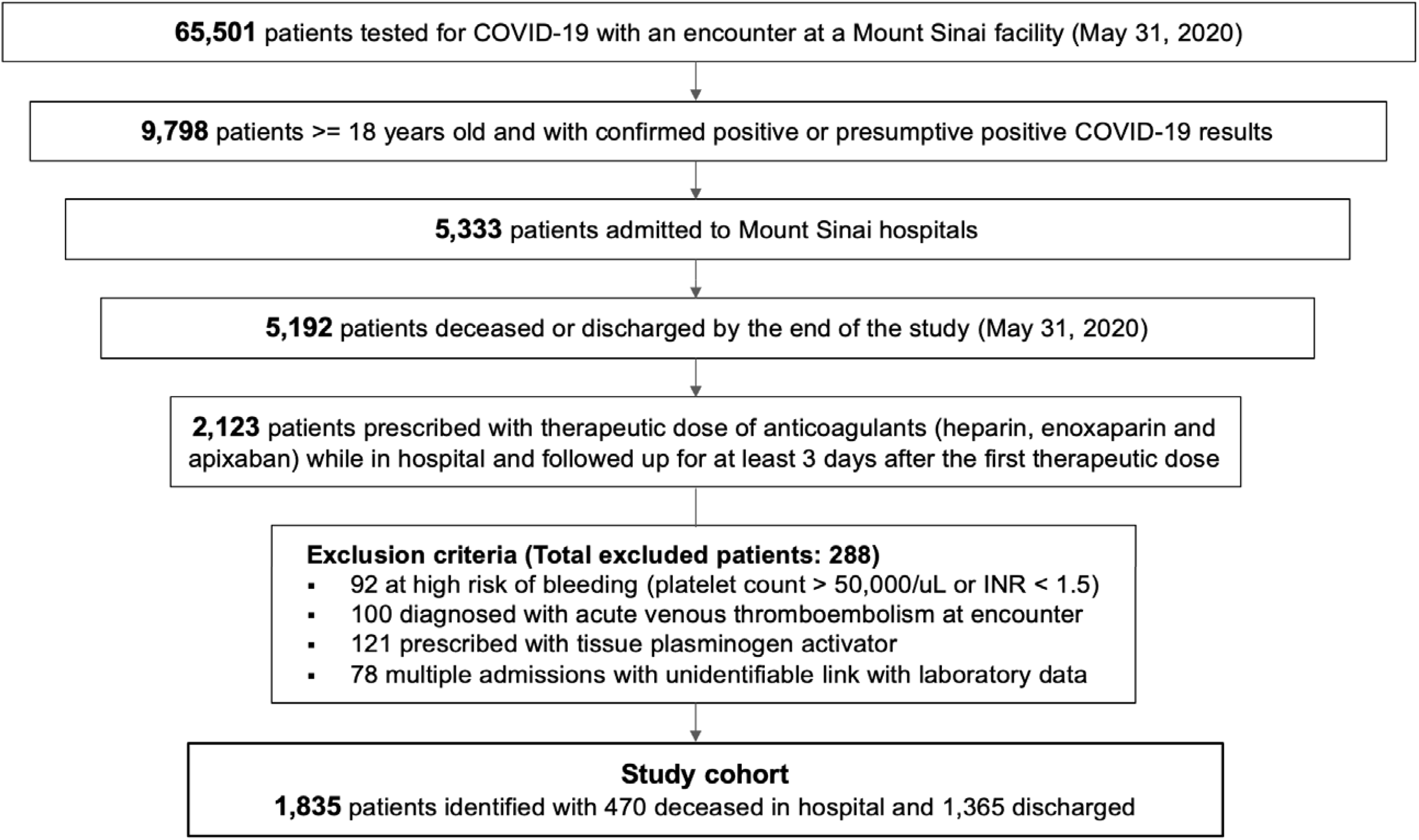
Selection of Study Cohort to Evaluate Role of Post-Anticoagulant D-Dimer as Predictive Biomarker for Mortality

### Post-anticoagulant D-dimer levels and COVID-19 mortality

Following therapeutic anticoagulation, the mean D-dimer was significantly higher for patients who died vs. those who were discharged from the hospital (median 3.71 ug/ml; [interquartile range, IQR 1.98, 8.05 ug/ml] vs. 1.69 ug/ml [IQR 0.86, 3.41 ug/ml], respectively; p<0.001). The difference in mean post-anticoagulant D-dimers between discharged vs. died groups was greater than the difference observed at baseline (2.02 ug/ml vs. 0.39 ug/ml, respectively; p<0.001). An increasing trend of post-anticoagulant D-dimers was observed for patients who died in the hospital (median slope: 0.09), while a decreasing trend was seen for those who were discharged (median slope: -0.05), with a significant difference between the changes in slope (p<0.001) (**Figure 2A**). The predictive power for in-hospital mortality of the logistic regression model with post-anticoagulant D-dimer mean level and its trend (AUC 0.76; 95% CI 0.74, 0.78) was significantly greater than the model with the baseline D-dimer (AUC 0.59; 95% CI 0.55, 0.61). Including baseline D-dimer levels to the model based on post-anticoagulant D-dimer did not further improve the prediction (AUC 0.76; 96%CI [0.74, 0.78]) (**Figure 2B**).

**Figure 2.**
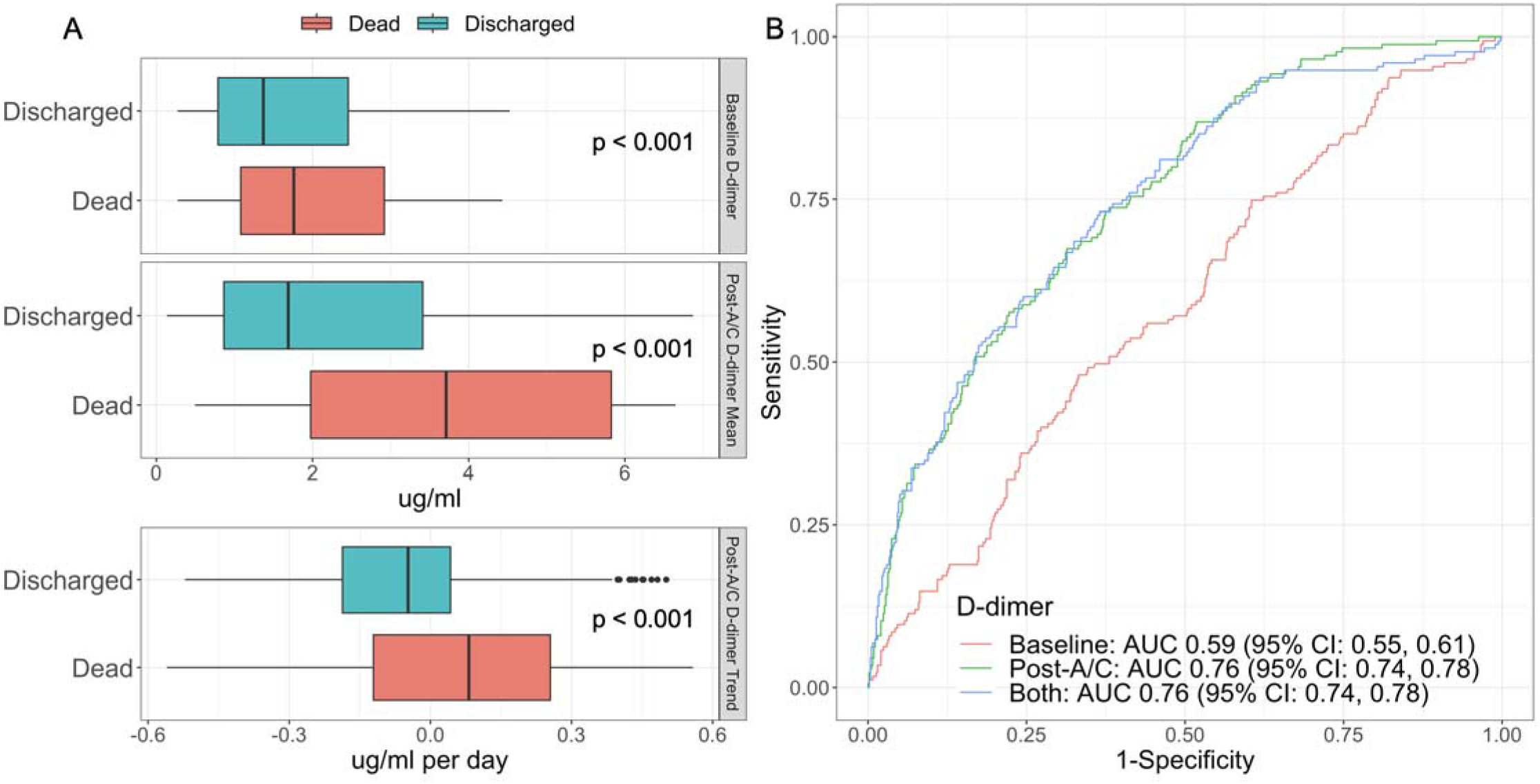
D-dimer distribution and its association with patient outcomes. (A) Boxplot of baseline and post-A/C D-dimer values. (B) ROCs of prediction models with baseline D-dimer, post-A/C D-dimer and both (AUCs 0.59, 0.76 and 0.76, respectively).

By stratifying the study cohort into four similar sized groups by combining high vs. low post-anticoagulant D-dimer means and increasing vs. decreasing trend, a significant difference was observed in the in-hospital mortality rates for patients within HI (49%) vs. HD (27%) vs. LI (21%) vs. LD (9%) groups (p<0.001) (**Figure 3A**). Patients with high mean post-anticoagulant D-dimer (>=2.5 ug/ml) were typically older, had more comorbidities, lower oxygen saturation, higher baseline D-dimers, higher leukocyte counts with lower lymphocyte percentages, worse kidney function and higher inflammatory markers than patients with low mean post-anticoagulant D-dimer (<2.5 ug/ml; p<0.001 for all comparisons) (**Supplementary Table 2**). Among patients within the high or low mean post-anticoagulant D-dimer groups, only lower baseline D-dimer was associated with increasing D-dimer trends (p<0.001) (**Figure 3B**).

**Figure 3.**
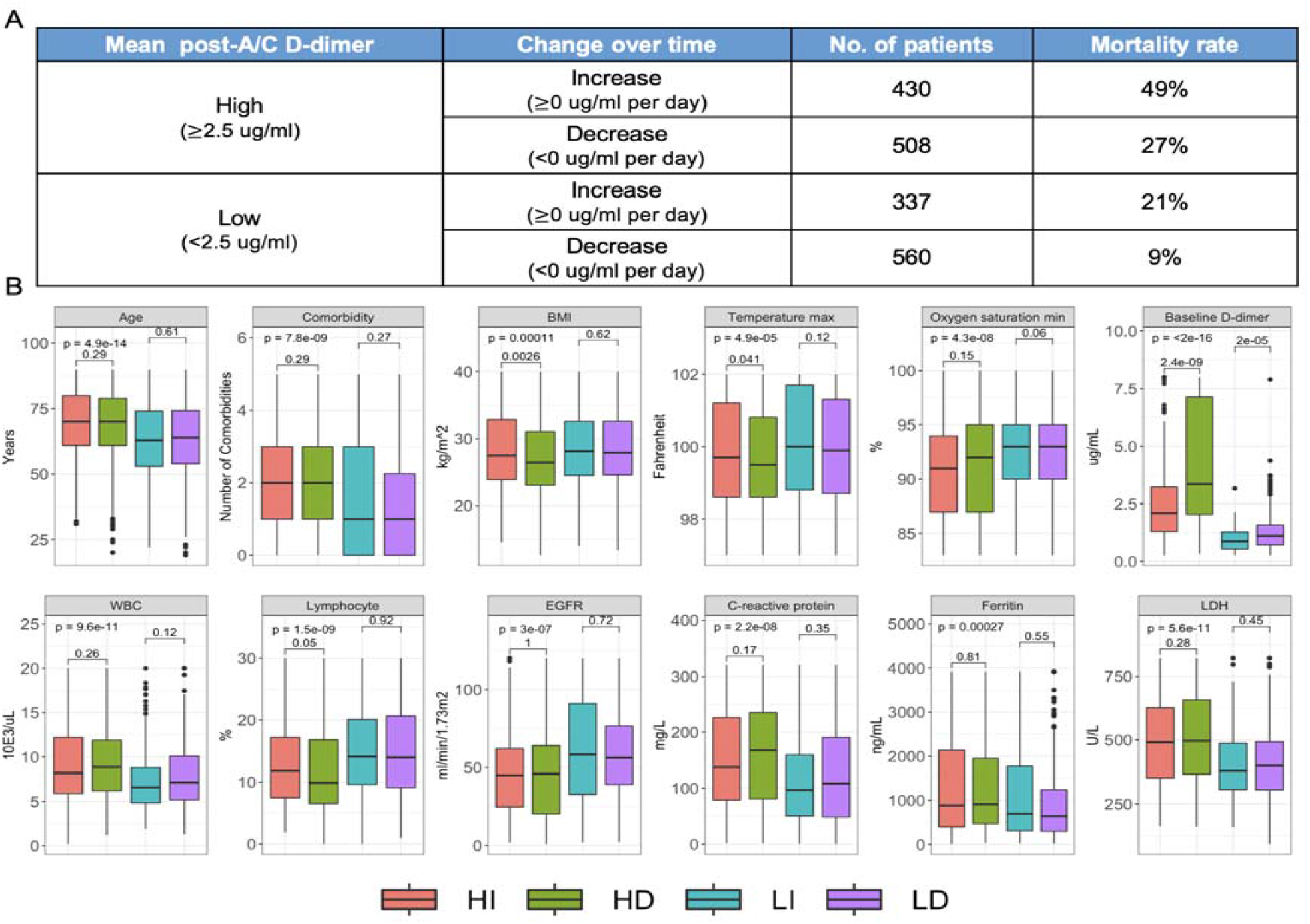
In-hospital mortality and baseline patient characteristics of four Post-A/C D-dimer groups. (A) In-hospital mortality rates by post-A/C D-dimer groups. (B) Baseline characteristics of patients with different D-dimer groups after A/C therapy.

### Post-anticoagulant D-dimer groups and in-hospital mortality

Jointly modeling post-anticoagulant D-dimer groups and 65 baseline covariates with penalized logistic regressions, 12 variables were selected to be predictive of in-hospital mortality through 10-fold cross validation based on the discovery subset. Among these, 10 variables were confirmed to be significantly associated with in-hospital mortality based on the validation subset (**Figure 4A)**. Compared to patients in the LD post-anticoagulant D-dimer group, patients in the HI post-anticoagulant D-dimer group were significantly the most likely to die during hospitalization (ORadj = 6.58; 95% CI [3.81,11.16]), followed by those in the LI post-anticoagulant D-dimer group (ORadj = 4.06; 95% CI [2.23, 7.38]) and HD group (ORadj = 2.37; 95% CI [1.37, 4.09]), after adjusting for the other pre-selected covariates. The post-anticoagulant D-dimer group was a stronger predictor of mortality than other covariates, such as acute kidney injury (ORadj = 1.99; 95% CI [1.34, 2.96]), or acute respiratory distress syndrome (ARDS; ORadj = 2.46; 95% CI 1.44-4.20) at admission (**Figure 4A)**. The baseline D-dimer value was not a significant predictor of mortality and was not selected in the final model.

**Figure 4.**
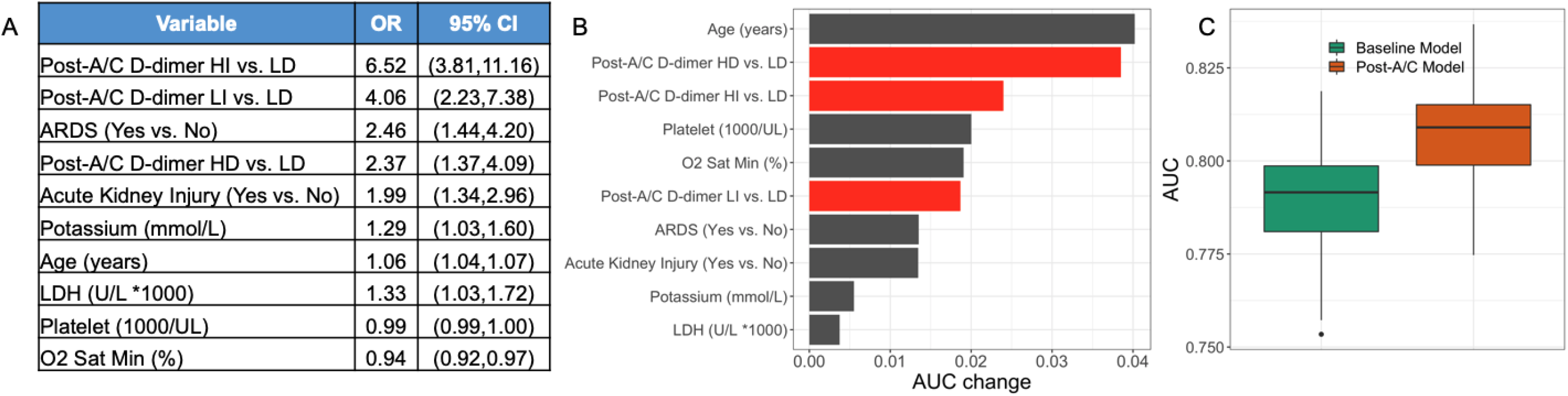
A multivariate prediction model for patients’ outcome. (A) ORs estimates, 95% CI for variables selected in the post-A/C model. (B) AUC differences between leave-one-predictor-out models and the full model for variables selected in the post-A/C model. (C) Comparison of 10-fold CV AUCs between baseline model and post-A/C model in 100 bootstrap samples.

### Robustness of the findings

When we evaluated the impact of individual predictors in the above model by calculating the change of the model’s AUC after excluding one variable at a time, age resulted in the largest AUC change (0.041), followed by HD post-anticoagulant D-dimer group (0.039), HI post-anticoagulant D-dimer group (0.024), platelet count (0.020), minimum oxygen saturation (0.020) and then LI post-anticoagulant D-dimer group (0.019) (**Figure 4B**). The combined effects of the four post-anticoagulant D-dimer groups had the greatest impact on model prediction (AUC change 0.054). Furthermore, when we compared the predictive powers of the above model based on post-anticoagulant D-dimer groups as well as the selected baseline variables with the predictive powers of a model based on only baseline variables, the AUCs of the post-anticoagulant D-dimer models are significantly higher than that of the baseline models based on 100 bootstrap data sets generated from the validation subset (**Figure 4C**).

## Discussion

In this retrospective study of 1,835 adult patients on therapeutic anticoagulation for thromboprophylaxis during admission for severe COVID-19 illness, we found high and independent predictive power of post-anticoagulant D-dimer levels for in-hospital mortality, while taking into consideration 65 other important covariates including patient demographics, comorbidities, vital signs and laboratory tests collected at baseline. We further identified patient trajectories of D-dimers values after anticoagulant therapy, which demonstrated significant differences in mortality rates and had the greatest impact on model prediction among all clinical characteristics under consideration. Therefore, post-anticoagulant D-dimer levels and trends are novel prognostic biomarkers that should be considered in the management of hospitalized COVID-19 patients.

Elevated D-dimer is among the most consistent markers of poor outcomes in COVID-19 patients. Several retrospective studies of hospitalized patients in Wuhan, China have demonstrated elevated D-dimer levels on admission with differing optimal cutoffs (starting at >0.5 ug/ml) to be predictive of in-hospital mortality^3,6,14,5^. However, while D-dimer at the time of admission may help stratify all COVID-19 patients according to mortality risk, it was not a significant predictor of mortality in our hospitalized cohort of COVID-19 patients with severe illness and receiving therapeutic anticoagulation for thromboprophylaxis. Instead, we found post-anticoagulant D-dimer levels to be highly predictive of in-hospital mortality in this group, with patients in the HI group 6.58 times more likely to die during the admission than patients in the LD group. Interestingly, patients in the LI post-anticoagulant D-dimer group had a higher risk of dying than those in the HD group, suggesting that the trend of the D-dimer following anticoagulation is more important than the three-day mean. As there were limited specific baseline patient characteristics associated with post-anticoagulant D-dimer trends, going forward it is critical that serial measurements are collected for more accurate prediction of in-hospital mortality.

While on anticoagulation, D-dimer and other coagulation parameters are commonly measured throughout the hospitalization. However, there is no consensus on how to incorporate such data to guide management decisions. Persistently elevated or rising D-dimer levels following anticoagulation may signify continued risk of large vessel and micro thrombotic events ^15,16^. Our findings provide a novel prognostic biomarker that can be widely incorporated into the treatment decision protocols for severe COVID-19 patients on therapeutic anticoagulation. We highlight an important subset of patients associated with especially poor outcomes (i.e., HI post-anticoagulant D-dimer) that can help guide resource allocation and prospective studies for emerging treatments. If proven effective in this setting, additional anticoagulants (i.e. antiplatelet therapy; TPA) may be considered to manage the potential worsening of coagulopathy in these patients. Conversely, future studies may explore if patients in the LD group can be changed to prophylactic doses of anticoagulant and whether they require continued thromboprophylaxis after hospital discharge.

### Limitations

There are limitations to this study worth discussing. Patients treated at a single tertiary hospital network in New York City may not be representative of the general population in the US and worldwide. There may be imprecisions of laboratory assays, which can alter the assessment of D-dimer. In addition, we were unable to account for unmeasured confounders that may affect D-dimer levels, a particular limitation inherent to all observational studies. Ongoing randomized controlled trials assessing the impact of therapeutic anticoagulation on COVID-19 outcomes should validate the ability of post-anticoagulant D-dimer levels to predict mortality. However, it may take significant time for the results of these trials to be reported. Therefore, our findings provide useful and immediate information to help guide management decisions in patients with severe COVID-19 illness.

## Conclusion

D-dimer levels and trends following initiation of anticoagulation have high and independent predictive value for in-hospital mortality for COVID-19 patients, and should be considered in management decisions for these patients.

## Data Availability

We did not generate new data, but used de-identified data from Mount Sinai Ware House, and the access of the data is regulated by Mount Sinai.

## Acknowledgement

We would like to express our sincerest condolences to the patients and their families who were affected by the COVID-19 outbreak. We greatly appreciate all the health care providers who contributed to the care of these patients. The Mount Sinai de-identified COVID-19 database was supported through the computational and data resources and staff expertise provided by Scientific Computing at the Icahn School of Medicine at Mount Sinai.

## Data Availability

The datasets analyzed during the current study are not publicly available due to United States Federal Health Insurance Portability and Accountability Act (HIPAA) compliance. A de-identified dataset may be available from the corresponding authors on reasonable request.

